# Factors associated with knowledge, attitudes, and behaviors regarding antiviral medications for COVID-19 among US adults

**DOI:** 10.1101/2023.12.11.23299148

**Authors:** E. Ivy Oyegun, Muyiwa Ategbole, Cynthia Jorgensen, Allison Fisher, Melissa Briggs Hagen, Lindsay Gutekunst, Eric Roberts, Emilia H. Koumans

**Author notes:** **Corresponding Author:** E. Ivy Oyegun, MPH, Epidemiology Branch, Coronavirus and Other Respiratory Viruses Division (CORVD), National Center for Immunization and Respiratory Diseases, Centers for Disease Control and Prevention. **Disclaimer**: The findings and conclusions are those of the authors and do not necessarily represent the official position of the Centers for Disease Control and Prevention.

## Abstract

**Background:** Little is known about public perceptions of antivirals for the treatment of mild-to-moderate COVID-19 in the United States (US). Our objective was to explore adult perceptions toward COVID-19 antivirals with the goal of improving outreach communications about antivirals for COVID-19.

**Methods:** During July 2022, potential respondents 18 years and older were randomly sampled from a national opt-in, non-representative, cross-sectional internet panel, with oversampling of African Americans, Hispanics, and adults 65 years and older. Respondents were asked about sociodemographic factors, and knowledge, attitudes, and perceptions regarding COVID-19 antivirals. Results were weighted to represent the non-institutionalized US adult population.

**Results:** Among 1,155 respondents, 51% were female, 60% were 18-49 years, 21% were 50-64 years, and 19% were 65 years or older. Compared to those aged 18-49 years and 50-64 years, a greater proportion of adults 65 years and older were knowledgeable about COVID-19 antivirals and would take them if they tested positive or their doctor recommended them. Adults 65 years and over and those reporting immunosuppression or disability had the highest rates of willingness to take antivirals. For all groups, the proportion of people willing to take antivirals increased by >20% if recommended by their doctor. Respondents in the 50-64 and 65+ groups who were sure they would take COVID-19 antivirals were more likely to be fully vaccinated and less likely to be living in isolation.

**Conclusion:** Groups that are less likely to have been vaccinated, those living in isolation, and those not sure about whether they would take an antiviral or not may be at risk for not receiving treatment to prevent severe COVID-19 outcomes. However, trust in doctor recommendations may be enough to overcome individual patient concerns about COVID-19 antivirals. Targeted initiatives to educate those at risk for severe COVID-19 outcomes about the effectiveness of antivirals, including those who are unvaccinated given their increased risk of severe disease, may be needed to further lower this population’s risk of severe COVID-19.

## Background and Introduction

FDA has authorized and the NIH treatment guidelines panel has recommended COVID-19 antivirals [1] for adults 50 years and over and for those with underlying medical conditions to reduce the risk for severe outcomes from COVID-19. However, among those eligible only a minority are receiving these medications. [2, 3] There are likely a multitude of reasons for this low uptake stemming from both provider and patient perspectives, as well as logistic considerations such as access. Potential contributing factors from the provider perspective for reduced use of antivirals include concerns about the risk of rebound as well as concerns about drug-drug interactions. [2, 4–7] While multiple articles have reported on disparities in COVID-19 treatment use, with significant differences reported by race/ethnicity and socio-economic status, there has been inadequate exploration of the reasons for these disparities or the interventions required to address them. [8, 9]

Public perception about the use of antivirals is poorly understood. Understanding psycho-social determinants of health such as knowledge, attitudes, and perceptions (KAPs) have proven useful in identifying potential underlying misconceptions and barriers hindering patient uptake of treatment, and in shaping targeted strategies for successful adoption and subsequently improved management of diseases. [10, 11] If knowledge about COVID-19 antivirals among those eligible is low, their full benefit may not be realized. [12] As such it is important to assess the underlying gaps in public knowledge, attitudes, and perceptions to better understand and identify opportunities and methods to improve antiviral uptake. Despite this, while literature exists on the public’s KAPs of COVID-19 stay-at-home orders, vaccinations, and boosters, there is limited literature on the public’s KAPs towards COVID-19 antivirals. [13, 14] The objective of this manuscript is to highlight KAPs as they relate to COVID-19 antiviral medications, focusing on adult awareness, attitudes, and perceived willingness to take the antivirals.

## Methods

### Sample

Between July 27, 2022 and July 31, 2022, KRC Research conducted an online cross-sectional survey of U.S. adults, ages 18 or older to understand attitudes and behaviors regarding antiviral medications for COVID-19. This survey was funded through a contract between the National Center for Immunizations and Respiratory Diseases Office of Communications and KRC Research, underwent IRB and Human Subjects Determination review and was determined to be non-research by CDC. The survey utilized non-probability sampling drawn from large national panels of participants who have access to the Internet and who opt-in to participate in periodic online sample surveys. Each survey was conducted in English. To ensure demographic representation for analyses, those who identified themselves as African American or Black, Hispanic, or were over the age of 65 were oversampled until at there were at least 200 respondents within each of those demographic segments. The completion rate of those who chose to participate was 85%. Results were weighted to represent the non-institutionalized US adult population using the 2020 census. [26]

### Survey

The following demographic variables were collected from each respondent, response options are included in parenthesis: age (free-text entry 18years and older included), race (White, Black or African-American, Asian, Other as a multiselect answer), ethnicity [Hispanic/Spanish/Latino descent] (yes, no), fully vaccinated status (yes, no), first and second booster status (yes, no), gender (male, female), education level (grade school or less, some high school, high school graduate, some college, 2-year college/technical school, 4-year college, some postgraduate work, postgraduate degree), state of residence (pre-filled drop down menu of all 50 states and D.C.), zip code (5-digit free text), residential community (urban community, suburban community, rural community), home ownership status (own, rent, live with others at no cost), number of adults and children in household (pre-filled drop down menu 0-10), employment status (work full-time, work part-time, self-employed, student, homemaker, retired, omitted, unemployed and seeking work, unemployed and not seeking work, unable to work because of a disability), income level (less than $25,000, $25,000-$29,999, $30,000-$34,999, $35,000-$39,999, $40,000-$49,999, $50,000-$59,999, $60,000-$74,999, $75,000-$99,999, $100,000-$124,999, $125,000-$149,999, $150,000-$199,999, $200,000 or more), political affiliation (Republican, Democrat, independent), immunocompromised status (yes, no, not sure). States were grouped according to the four census regions: Northeast, Midwest, South, West. In-depth questions about underlying medical conditions were not asked.

### Definitions

We stratified the study population into four mutually exclusive groups based on age, immunosuppression, and disability for the outcome analysis. These groups were: adults 18-49 years with no immunosuppression or disability (18-49), adults 50-64 years with no immunosuppression or disability (50-64), adults 65 years and over with no immunosuppression or disability (65+), and adults aged 18 and older who have immunosuppression or disability (Immunosuppressed or Disabled).

### Outcomes

Respondents were asked to select their level of agreement with questions measuring their knowledge, attitudes, perceptions, preferences, and intentions about antivirals. For each question, the respondent was able to select a variety of graduated responses. Survey questions analyzed for this manuscript contributed to one of three primary outcomes: 1) knowledge of antivirals, 2) attitudes towards antivirals, and 3) perceived willingness to take antivirals. Each of the two questions for outcomes 2 and 3 were analyzed separately.

The first outcome, *Knowledge of antivirals,* was measured via the following question “Have you heard of antiviral medications to treat people who get infected with COVID-19?” The response options for this question were “Yes”, “No”, “Not sure”. Discriminant analysis was conducted to identify the characteristics that best predicted knowledge of antivirals and to identify the largest differences between those who were knowledgeable versus those who were not. The “No” and “Not Sure” responses were combined, and the “Yes” response was used as the reference group.

The second outcome, *Attitudes towards antivirals,* was measured via responses to the following two questions “New antiviral medications are effective in decreasing the seriousness of symptoms from a COVID-19 infection” and “New antiviral medications are effective in reducing a person’s chance of getting hospitalized or dying from COVID-19”. Response options for each question were Strongly agree, Agree, Neither agree nor disagree, Disagree, Strongly disagree. Multiple linear regression was used to identify the characteristics that best predicted attitudes towards antivirals and to identify the largest differences between those who expressed belief in efficacy of antivirals vs those who did not, with “Strongly agree” or “Agree” as the reference group.

The last outcome, *Perceived willingness to take antivirals,* was measured via responses to the following two questions “I would take antiviral medications if I tested positive for COVID-19.” and “I would take antiviral medications for a COVID-19 infection if my doctor recommended it” (response options for both questions = Yes, No, Not Sure). Discriminant analysis was used to identify the characteristics that best predicted perceived willingness to take antivirals and to identify the largest differences between those who were willing versus those who were not willing. Responses were modelled as “Yes” vs “Not Sure”, with “Yes” as the reference group. The “No” response was not included in the analysis to focus on differences between respondents who were willing vs those whose opinion was not fixed against antivirals.

Results include beta coefficients (β) which represent how strongly each variable predicted its corresponding outcome. Higher coefficients represent higher predictive strength and vice versa. The + sign indicates that respondents with that characteristic where more likely to agree with the corresponding outcome statement, while the - sign indicates that respondents with that characteristic where less likely to agree with the corresponding outcome statement.

The strength of each characteristic’s predictability may vary for each outcome. Therefore, to ensure each outcome has multiple options for communication strategies while focusing on the most predictive characteristics, results will highlight the three best predicting characteristics for each outcome (knowledge, attitudes, and willingness) above a β = +/- 0.2.

#### Statistical analyses

Since the goal of the data collection and analysis was to generate general information for communications activities such as which subgroup to target due to higher or lower knowledge, attitude, or willingness, precision about differences between subgroups in each demographic category were not prioritized. Therefore, multiple linear regression and discriminant analysis were used to identify which three demographic characteristics *best* predicted respondents who agreed with each of outcomes. Results between linear regression/discriminant analyses and logistic regression were extremely similar as can sometimes occur, therefore choosing between each approach can come down to stylistic preferences. [25] In this case linear regression/discriminant analysis was chosen because the format of its output was more aligned with the study’s objective.

## Results

### Demographics

The study population consisted of 1,155 adult respondents aged 18 years and older. Table 1 displays characteristics of respondents by age and risk groups. Of the respondents, 588 (51%) were female, 692 (60%) were 18-49 years, 240 (21%) were 50-64 years, and 223 (19%) were aged 65 years or older.

**Table 1.** ^a, f^ Characteristics of adult survey respondents about knowledge, attitudes, and willingness to take COVID-19 antivirals, weighted %, by age group and immunosuppressed or disability status, US, July 2022

|  |  | Non-Immunosuppressed or non-Disabled |  |  | Immunosuppressed or Disabled |
| --- | --- | --- | --- | --- | --- |
|  |  | 18-49<br>(n=421)<br>% | 50-64<br>(n=124)<br>% | 65+<br>(n=115)<br>% | Immunosuppressed or Disabled<br>(n=366)<br>% |
| Total |  | 36.0 | 11.0 | 10.0 | 32.0 |
| Gender | Women | 52.0 | 45.0 | 47.0 | 43.0 |
|  | Men | 47.0 | 55.0 | 53.0 | 57.0 |
| Age (years) | 18-24 | 22.0 | 0.0 | 0.0 | 6.0 |
|  | 25-34 | 33.0 | 0.0 | 0.0 | 15.0 |
|  | 35-44 | 33.0 | 0.0 | 0.0 | 13.0 |
|  | 45-54 | 13.0 | 35.0 | 0.0 | 17.0 |
|  | 55-64 | 0.0 | 65.0 | 0.0 | 22.0 |
|  | 65 or older | 0.0 | 0.0 | 100.0 | 27.0 |
| Region<sup>b</sup> | Northeast | 16.0 | 21.0 | 18.0 | 18.0 |
|  | Midwest | 22.0 | 18.0 | 19.0 | 22.0 |
|  | South | 37.0 | 36.0 | 42.0 | 37.0 |
|  | West | 25.0 | 25.0 | 21.0 | 23.0 |
| Race / Ethnicity | Non-Hispanic White | 65.0 | 75.0 | 85.0 | 79.0 |
|  | Non-Hispanic Black | 20.0 | 10.0 | 9.0 | 12.0 |
|  | Hispanic | 22.0 | 15.0 | 4.0 | 16.0 |
| Community type | Urban | 36.0 | 17.0 | 20.0 | 35.0 |
|  | Suburban | 45.0 | 61.0 | 57.0 | 41.0 |
|  | Rural | 19.0 | 21.0 | 23.0 | 24.0 |
| Marital Status | Married | 34.0 | 50.0 | 65.0 | 41.0 |
|  | Not married | 66.0 | 50.0 | 35.0 | 59.0 |
| Children in home | Children | 49.0 | 22.0 | 3.0 | 29.0 |
|  | No children | 51.0 | 78.0 | 97.0 | 71.0 |
| Education<sup>c</sup> | High school or less | 39.0 | 35.0 | 27.0 | 42.0 |
|  | Some college/trade | 24.0 | 26.0 | 26.0 | 27.0 |
|  | College grad or more | 38.0 | 40.0 | 47.0 | 31.0 |
| Employment status<sup>d</sup> | Employed | 74.0 | 68.0 | 16.0 | 43.0 |
|  | Retired | 1.0 | 15.0 | 81.0 | 29.0 |
|  | Student | 5.0 | 0.0 | 0.0 | 1.0 |
|  | Homemaker | 7.0 | 10.0 | 1.0 | 4.0 |
|  | Not employed<br>currently/unable to<br>work | 11.0 | 1.0 | 0.0 | 27.0 |
| Home ownership | Own | 41.0 | 69.0 | 89.0 | 49.0 |
|  | Rent | 48.0 | 31.0 | 11.0 | 45.0 |
|  | Live with others at no<br>cost | 11.0 | 1.0 | 0.0 | 5.0 |
| Political Party | Democrats & Ind.<br>Leaners | 51.0 | 48.0 | 49.0 | 48.0 |
|  | Republicans & Ind.<br>Leaners | 31.0 | 36.0 | 40.0 | 32.0 |
|  | Independent/others do<br>not lean | 17.0 | 15.0 | 11.0 | 19.0 |
| Household Income<sup>e</sup> | Less than \$35k | 34.0 | 30.0 | 25.0 | 46.0 |
| | \$35k to less than \$50k | 13.0 | 14.0 | 14.0 | 10.0 |
| | \$50k to less than 75k | 18.0 | 16.0 | 18.0 | 17.0 |
| | \$75k to less than \$100k | 12.0 | 11.0 | 14.0 | 5.0 |
| | \$100k or more | 23.0 | 29.0 | 29.0 | 22.0 |
| Immunocompromised | Yes | 0.0 | 0.0 | 0.0 | 100.0 |
|  | No | 46.0 | 18.0 | 15.0 | 21.0 |
| Serious Health Problem | Yes | 0.0 | 0.0 | 0.0 | 100.0 |
|  | No | 53.0 | 21.0 | 18.0 | 8.0 |
| Vaccination status | Fully vaccinated | 21.0 | 26.0 | 28.0 | 25.0 |
|  | Received First Booster | 29.0 | 15.0 | 17.0 | 40.0 |
|  | Received Second Booster | 0.0 | 15.0 | 26.0 | 59.0 |

a Column percentages are displayed here; Only includes respondents with complete demographic data
b The 50 states and D.C. were grouped into these 4 Census regions
c Education levels were collapsed as follows: High School or less= Grade school or less, Some high school, High school graduate; Some college/trade = Some college, 2-year college/technical school; College graduate or more= 4-year college, Some postgraduate work, Postgraduate degree
d Employed includes: Work full-time, Work part-time, and Self-employed; Not employed currently/unable to work includes: Unemployed and seeking work, Unemployed and not seeking work, Unable to work because of a disability
e Income levels were collapsed as follows: Less than \$35k includes Less than \$25,000, \$25,000-\$29,999, and \$30,000-\$34,999; \$35k to less than \$50k includes \$35,000-\$39,999 and \$40,000-\$49,999; \$50k to less than \$75k includes \$50,000-\$59,999 and \$60,000-\$74,999; \$75k to less than \$100k includes \$75,000-\$99,999; \$100k or more includes \$100,000-\$124,999, \$125,000-\$149,999, \$150,000-\$199,999, and \$200,000 or more
f Results were weighted to represent the non-institutionalized US adult population using the 2020 census

### Perceptions and willingness to take antivirals

The following results are shown in Figure 1 which summarizes knowledge and attitudes and perceptions about antiviral effectiveness and willingness to take them by age and risk group. Respondents 65 years and older represented the highest proportion (69%) who had heard of antiviral medications, followed by the Immunosuppressed or Disabled group (67%). A greater proportion of respondents 65 years and older agreed that antivirals reduced the chances of developing severe outcomes such as serious symptoms (78%) or of being hospitalized/dying from COVID-19 (75%) compared to those aged 50-64 (64%, 63% respectively) and those aged 18-49 (52%, 52% respectively). This was slightly higher than among respondents that were immunosuppressed or disabled (74%, 69% respectively). When asked about willingness to take antivirals, a greater proportion of respondents 65 years and older would take antivirals if they tested positive for COVID-19 (64%), or if their doctor recommended it (93%) compared to those aged 50-64 (53%, 82% respectively) and those aged 18-49 (50%, 70% respectively); this was again slightly higher than responses among those who were immunosuppressed or disabled (63%, 88% respectively).

**Figure 1.**
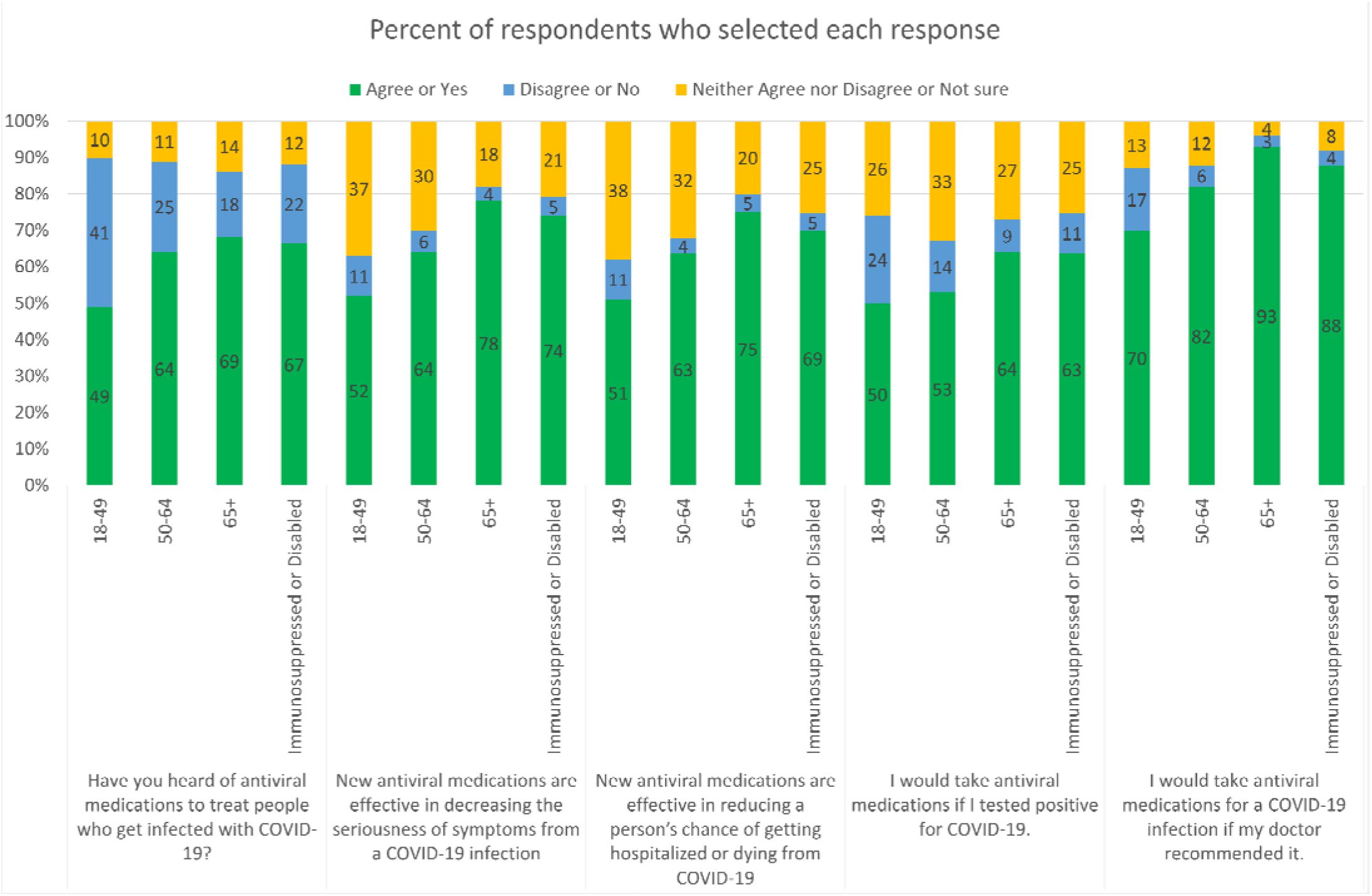
Proportion of respondents responding strongly agree/agree or yes (green), neither agree nor disagree or not sure (yellow), or disagree or no (blue) to question about antiviral knowledge, attitudes toward antivirals, and willingness to take antivirals.

The results below highlight the three best predicting characteristics for each outcome (knowledge, attitudes, and willingness) above β = +/- 0.2. Additional data on model characteristics is presented in Tables 2a, 2b, and 2c.

**Table 2a:** Characteristics associated with knowledge of antivirals among participants 18-49 years old, 50-64 years old, 65 and older, and those with disability or immunosuppression (any age): YES vs NO+NOT SURE

|  | 18-49<br>(N=421) | 50-64<br>(N=124) | 65+<br>(N=115) | Immunosuppressed<br>or Disabled<br>(N=366) |
| --- | --- | --- | --- | --- |
| Estimated percent of variance accounted for by the model | 9.3% | 15.1% | 38.7% | 14.9% |
| Characteristic* | Statistic ($\beta$) | Statistic ($\beta$) | Statistic ($\beta$) | Statistic ($\beta$) |
| Education | 0.484 | - | 0.632 | 0.419 |
| Hispanic | - | - | 0.432 | -0.413 |
| Household income | 0.551 | - | 0.591 | - |
| Separated, divorced, widowed | - | -0.508 | - | -0.273 |
| Age | 0.44 | - | - | - |
| Boosted | - | - | - | 0.374 |
| Household size | - | - | - | 0.274 |
| Male | - | - | -0.7 | - |
| Northeast region | - | 0.66 | - | - |
| Not working | - | - | - | -0.423 |
| Rural | - | -0.524 | - | - |
| South | - | - | -0.581 | - |

\*Discriminant analysis was conducted to identify the characteristics from Table 1 that best predicted knowledge of antivirals and to identify the largest differences between those who were knowledgeable versus those who were not. The “No” and “Not Sure” responses were combined, and the “Yes” response was used as the reference outcome. The referent groups for income = \$57,000 +, for education = 2year college +, and for age = 55years + (35years + in the 18-49 group). The referent groups for all other independent characteristics was not having that characteristic, e.g. referent group of “boosted” is “not boosted”. Displayed characteristics account for strongest predictive model. All $\beta$ values are significant at $p < 0.05$ or less “-” = Not Significant
The beta coefficients ( $\beta$ ) reported represent how strongly each variable predicted this outcome, while +/- indicates the likelihood that respondents who answered Yes to this outcome were more or less likely (respectively) to have the characteristic variable compared to those who answered Not Sure.

**Table 2b:** Characteristics associated with attitudes toward antivirals among 18-49 year olds, 50-64 year olds, 65 and older, and those with disability or immunosuppression (any age): 5 point Likert scale from Strongly Agree to Strongly Disagree.

| New antiviral medications are effective in decreasing the seriousness of symptoms from a COVID-19 infection: STRONGLY AGREE/AGREE as reference |  |  |  |  |
| --- | --- | --- | --- | --- |
|  | 18-49<br>(n=374) | 50-64<br>(n=144) | 65+<br>(n=127) | Immunosuppressed<br>or Disabled<br>(n=381) |
| Estimated percent of variance accounted for | 8.3% | 13.3% | 25.5% | 16.2% |
| Characteristic* | Statistic ($\beta$) | Statistic ($\beta$) | Statistic ($\beta$) | Statistic ($\beta$) |
| Boosted | - | - | 0.355 | 0.155 |
| Age | 0.168 | - | - | - |
| Children 7 to 12 | - | 0.166 | - | - |
| Double boosted | - | 0.19 | - | - |
| Education | - | - | - | 0.235 |
| Fully vaccinated | 0.166 | - | - | - |
| Income | - | -0.266 | - | - |
| Male | - | - | -0.259 | - |
| Northeast region | - | 0.184 | - | - |
| Not working | - | - | - | -0.113 |
| Political party or lean | -0.117 | - | - | - |
| Rent a home | -0.125 | - | - | - |
| Retired | - | - | - | 0.122 |
| Single | - | 0.207 | - | - |
| Urban | - | - | -0.19 | - |
| New antiviral medications are effective in reducing a person's chance of getting hospitalized or dying from COVID-19: STRONGLY AGREE/AGREE as reference |  |  |  |  |
|  | 18-49 no risk<br>(n=373) | 50-64 no risk<br>(n=145) | 65+ no risk<br>(n=127) | Immunosuppressed<br>or Disabled<br>(n=381) |
| Estimated percent of variance accounted for | 8.8% | 21% | 36.3% | 13.3% |
| Characteristic* | Statistic ($\beta$) | Statistic ($\beta$) | Statistic ($\beta$) | Statistic ($\beta$) |
| Boosted | 0.224 | 0.204 | 0.591 | 0.256 |
| Age | 0.166 | 0.39 | - | 0.126 |
| Education | - | - | 0.222 | 0.144 |
| Male | - | - | -0.198 | -0.132 |
| Political party or lean | -0.12 | -0.273 | - | - |
| Children 7 to 12 in household | - | 0.201 | - | - |

\*Multiple linear regression was used to identify the characteristics that best predicted attitudes towards antivirals and to identify the largest differences between those who expressed belief in efficacy of antivirals vs those who did not, with “Strongly agree” or “Agree” as the outcome reference group. The referent groups for income = \$5 7,000 +, for education = 2year college +, and for age = 55years + (35years + in the 18-49 group). The referent groups for all other independent characteristics was not having that characteristic, e.g. referent group of “boosted” is “not boosted”. Displayed characteristics account for strongest predictive model. All $\beta$ values are significant at $p < 0.05$ or less
“-” = not significant
The beta coefficients ( $\beta$ ) reported represent how strongly each variable predicted this outcome, while +/- indicates the likelihood that respondents who answered Strongly Agree to this outcome were more or less likely (respectively) to have the characteristic variable compared to those who did not answer Strongly Agree.

**Table 2c:** Characteristics associated with willingness to take antivirals among 18-49 year olds, 50-64 year olds, 65 years and older, and those with disability or immunosuppression (any age): Yes vs Not Sure.

| Predicted willingness to take anti-viral if test positive: YES (reference) vs NOT SURE |  |  |  |  |
| --- | --- | --- | --- | --- |
|  | 18-49 (n=313) | 50-64 (n=103) | 65+ (n=101) | Immunosuppressed<br>or Disabled<br>(n=315) |
| Estimated percent of<br>variance accounted for | 6.8% | 15.9% | 8.5% | 12.7% |
| **Characteristic\*** | **Statistic ($\beta$)** | **Statistic ($\beta$)** | **Statistic ($\beta$)** | **Statistic ($\beta$)** |
| Boosted | 0.606 | - | - | 0.783 |
| Separated, divorced,<br>widowed | - | -0.596 | - | -0.504 |
| Double boosted | - | 0.647 | - | - |
| Education | - | - | 0.7 | - |
| Hispanic | 0.507 | - | - | - |
| Income | - | -0.703 | - | - |
| Living at no cost | - | - | - | -0.373 |
| Own a home | 0.722 | - | - | - |
| Western region | - | -0.47 | - | - |
| Work full time | - | - | 0.699 | - |
| Predicted willingness to take anti-viral if doctor recommended: YES (reference) vs NOT SURE |  |  |  |  |
|  | 18-49 no risk<br>(n=341) | 50-64 no risk<br>(n=115) | 65+ no risk<br>(n=109) | Immunosuppressed<br>or Disabled<br>(n=338) |
| Estimated percent of<br>variance accounted for | 11% | 16.1% | 37.3% | 15.9% |
| **Characteristic\*** | **Statistic ($\beta$)** | **Statistic ($\beta$)** | **Statistic ($\beta$)** | **Statistic ($\beta$)** |
| Fully vaccinated | 0.442 | 0.719 | - | 0.474 |
| Boosted | - | - | -0.471 | 0.413 |
| Not working | - | - | 0.323 | -0.467 |
| Southern region | 0.563 | -0.605 | - | - |
| Children under 7 in<br>household | - | - | 0.718 | - |
| Hispanic ethnicity | 0.496 | - | - | - |
| Household size | - | - | 0.347 | - |
| Own a home | 0.542 | - | - | - |
| Single | - | - | 0.508 | - |
| Work Part Time or self-employed | 0.388 | - | - | - |

\*Discriminant analysis was used to identify the characteristics that best predicted perceived willingness to take antivirals and to identify the largest differences between those who were willing versus those who were not willing. Responses were modelled as “Yes” vs “Not Sure”, with “Yes” as the reference group. The referent groups for income = \$57,000 +, for education = 2year college +, and for age = 55years + (35years + in the 18-49 group). The referent groups for all other independent characteristics was not having that characteristic, e.g. referent group of “boosted” is “not boosted”. Displayed characteristics account for strongest predictive model. All $\beta$ values are significant at $p < 0.05$ or less
Footnote: “-” = Not Significant
The beta coefficients ( $\beta$ ) reported represent how strongly each variable predicted this outcome, while +/- indicates the likelihood that respondents who answered Yes to this outcome were more or less likely (respectively) to have the characteristic variable compared to those who answered Not Sure.

### Results by group

#### Immunosuppressed or Disabled Group

In the Immunosuppressed or Disabled group, knowledgeable respondents were more likely to have more education (β=0.419), and less likely to be non-working (β= -0.423), or Hispanic (β= -0.413) (Table 2a). Respondents who strongly agreed that COVID-19 antivirals could reduce symptom severity were more likely to be better educated (β= 0.235), while respondents who strongly agreed that COVID-19 antivirals could reduce risk of hospitalization or death were more likely to be boosted (β= 0.256) (Table 2b).

Respondents who were willing to use antivirals if they tested positive for COVID-19 were more likely to be boosted (β = 0.783) and less likely to be separated, divorced, or widowed (β = -0.504) or to live with others at no cost (β = - 0.373). Conversely, respondents who were willing to use antivirals if their doctor recommended it were more likely to be fully vaccinated (β= 0.474), or boosted (β= 0.413), and less likely to be unemployed (β= -0.467) (Table 2c).

### 65 years and older group

Knowledgeable respondents in this group were more likely to be educated (β= 0.632) or have higher incomes (β= 0.591). They were also less likely to be male (β= -0.7) (Table 2a).

Respondents who strongly agreed that COVID-19 antivirals could reduce symptom severity were more likely to be boosted (β= 0.355), and less likely to be male (β= -0.259). While respondents who strongly agreed that COVID-19 antivirals could reduce risk of hospitalization or death were more likely to be boosted (β=0.591), or older (β= 0.222) (Table 2b).

Respondents who were willing to use antivirals if they tested positive for COVID-19 were more likely to be more educated (β= 0.7) or work full time (β=0.699). While respondents who were willing to use antivirals if their doctor recommended it were more likely to be fully vaccinated (β= 0.817), or have higher incomes (β= 0.373), and less likely to live with fewer adults (β= -0.587)) (Table 2c).

### 50-64 years group

Knowledgeable respondents in the 50-64 years age group were more likely to live in the Northeast region (β= 0.66), and less likely to live in a rural setting (β= -0.524), or to be separated, divorced, or widowed (β= 0.508) (Table 2a).

Respondents who strongly agreed that COVID-19 antivirals could reduce symptom severity were more likely to have lower incomes (β= -0.266) or be single (β=0.207). While respondents who strongly agreed that COVID-19 antivirals could reduce risk of hospitalization or death were more likely to be older (β= 0.39), have Democratic leanings (β= -0.273), or boosted (β=0.204) (Table 2b).

Respondents who were willing to use antivirals if they tested positive for COVID-19 were more likely to have lower income (β= -0.703), or be double boosted (β=0.647), and less likely to be separated, divorced, or widowed (β= -0.596). While respondents who were willing to use antivirals if their doctor recommended it were more likely to be fully vaccinated (β= 0.719), and less likely to live in the Southern region (β= -0.605) (Table 2c).

### Thematic Results

Compared to lower level of education, higher level of education was a strong predictor of knowledge in the 65 and older, and Immunosuppressed or Disabled groups (β= +0.632, +0.419 respectively) with knowledgeable respondents more (+β) likely to be higher educated. Marital status was predictive of knowledge among the 50-64 age group and the Immunosuppressed or Disabled group (β = -0.508, -0.273 respectively), with knowledgeable respondents less (-β) likely to be separated, divorced, or widowed compared to those who were married.

In all study groups, compared to not being boosted, being boosted was a predictor of the belief that antivirals lessen the chance of severe symptoms, hospitalization, or death: 50-64 (β = +0.204), 65+ (β = +0.591), and Immunosuppressed or Disabled (β = +0.256). Marital status was predictive of willingness to take antivirals if a COVID-19 test is positive for the 50-64 (β= -0.596) and the Immunosuppressed or Disabled groups (β= -0.504), with willing respondents less likely (-β) to be separated, divorced, or widowed compared to those who were married.

Compared to not being fully vaccinated, being fully vaccinated was strongly predictive of willingness to take antivirals if their doctor recommended it for all groups except those aged 65 and older: 50-64 (β= +0.719), Immunosuppressed or Disabled group (β= +0.474), with willing respondents more likely to be fully vaccinated (+β).

## Discussion

To our knowledge, this is one of the first reports of public knowledge, attitudes, and willingness to use antivirals to reduce risk for severe COVID-19 in the United States. This cross-sectional survey, performed approximately 6 months after antivirals became available in the U.S., had diverse representation from each HHS region in the U.S., and was weighted to the non-institutionalized U.S. Census adult population. The proportion of participants willing to take antivirals if recommended was highest (93%) among those aged more than 65 years and those who were immunosuppressed or disabled (88%) and was associated with having received a booster among the latter. While characteristics that were associated with knowledge, attitudes, and willingness to take antivirals differed by age and risk groups, some of these characteristics overlapped. For example, among most groups, higher education was associated with knowledge of antivirals, and among all groups having had vaccinations or a booster was associated with attitudes toward and willingness to use antivirals. These findings have important implications for outreach efforts and communication about COVID-19 antivirals by providing evidence on how to target efforts, for example towards those with lower education and who are unvaccinated, to improve antiviral knowledge and uptake.

### Knowledge

People who are most at risk for severe COVID-19 outcomes, those older than age 65 and those who had a disability or were immunosuppressed, were more likely to know about antivirals if they reported higher levels of income and education. These findings about antiviral knowledge are similar to earlier studies examining knowledge of COVID-19 more generally; these studies also found that socio-demographics, including income and education, were associated with more accurate knowledge of COVID-19 and those with lower education or income levels had less knowledge and were less accurate. [15, 16] This suggests that characteristics affecting knowledge which were noted earlier in the pandemic also affect knowledge about antivirals, and that educational efforts about antivirals and risk have been more successful among those with a higher income and education. [17] These findings support the need for additional efforts to inform those with lower income and level of education about COVID-19 risk and about antivirals that address health literacy concerns raised early in the pandemic, and will continue to be relevant as new sub-variants arise. [18]

Additional characteristics, such as being separated, divorced, or widowed was associated with being less likely to have COVID-19 antiviral knowledge, while living with other adults and being boosted, were associated with increased knowledge among those at higher risk. These findings suggest that living alone or social isolation is one factor that may negatively affect knowledge [19].

### Attitudes towards antivirals

In all groups examined, we found that having been vaccinated was associated with the beliefs that antivirals reduce the severity of symptoms and that antivirals reduce the likelihood of hospitalization or death. These findings, in combination with the associations of income and education with antiviral knowledge, are similar to studies examining factors associated with vaccination; where people with higher incomes and higher levels of education are more likely to believe in the efficacy of pharmaceuticals and more likely to report acceptance if offered [21].

Other characteristics associated with having a positive attitude toward antivirals preventing hospitalization were older age (within each of the, 50-64, 65+, and Immunosuppression or Disability groups), democratic-leaning political affiliation (in the 50-64 group), and female gender (in the 65+ and Immunosuppression or Disabled group). Since the beginning of the pandemic, educational efforts have emphasized that people of older age are at higher risk for severe illness; this finding represents an achievement in risk communication. The finding that political affiliation is associated with attitudes toward antivirals is similar to the finding about attitudes around vaccination [22], where democratic political affiliation has been associated with willingness to get vaccinated. Female gender also played a role in attitudes toward antivirals for those 65 and older and for those who were immunosuppressed or disabled. The findings around political affiliation and gender may represent opportunities for distinct efforts in further communications and evaluations by focusing informational and outreach efforts towards males and those who are not democratic-leaning to improve COVID-19 antiviral uptake.

Furthermore, within the 50-64 and 65 and older groups, the largest proportion of respondents who selected Not sure or Neither agree nor Disagree was in response to knowledge of COVID-19 antivirals and taking antivirals if they tested positive. Among the 65 and older group, these respondents tended to be less likely to have higher education or to work full time. Among the 50-64 group, they tended to have higher incomes or to be less likely to be double boosted, while among the 50-64 and the group with immunosuppression or disability, they were less likely to be separated, divorced, or widowed. Additionally, the 12% of respondents in the 50-64 group who were not sure if they would take antivirals if their doctor recommended it were more likely to live in the Southern region and less likely to be fully vaccinated. These characteristics describe subpopulations of at-risk groups who could be targeted and who might benefit from specific messages to address perceptions about COVID-19 antivirals and possibly willingness to use them. Since characteristics vary by community, identifying multiple characteristics can be beneficial by allowing local entities and communities to identify local prevalent characteristics and focus their messaging and resources.

While some proportion of respondents in each age group were unsure about the effectiveness of antivirals and were neutral about taking them if they tested positive for COVID-19, far fewer respondents within each age group were unsure about taking antivirals if their doctor prescribed them. This reiterates prior findings that, despite personal attitudes and beliefs about health options, doctor’s recommendations are a potentially trusted source of information likely capable of overcoming individual patient concerns. As such, this presents an opportunity for targeted interventions such as messaging and providing doctors with tools to strengthen the antiviral recommendations to at-risk groups of concern, and possibly increase antiviral use among at-risk groups for whom they are recommended.

### Perceived willingness to take antivirals

Willingness to take antivirals given a positive test was associated with being boosted or double boosted among 50-54 years, and those with immunosuppression or disability, findings similar to the characteristics associated with knowledge and attitudes toward antivirals. [10] Other socioeconomic factors indicating higher level of resources, consistent with the factors associated with knowledge and attitude, were also associated with willingness to take antivirals, such as working full time, owning a home, and education. Except among those older than 65 years, being fully vaccinated or boosted were also associated with being willing to take an antiviral if a doctor recommended it in each group.

Antivirals can reduce the risk of hospitalization, post-COVID conditions or Long COVID, and death [23], but less than 50% of those eligible in studies examining electronic medical record data are prescribed them [3], therefore the full potential life-saving use of antivirals is currently not being met. Provider-level barriers have been described [2], but less is understood about knowledge, attitudes, and perceptions of people who might get COVID-19 and might be eligible for treatment. This study illuminates some of the factors that are associated with knowledge of, attitudes toward, and willingness to take antivirals. While certain groups of people appear to have adequate knowledge and importantly, some are willing to take an antiviral on the advice of a doctor, the consistent association with this willingness was previous vaccination.

## Strengths and Limitations

The strengths of this study were the randomly selected participants, the oversampling for under-represented populations, the weighting to account for the U.S. population, and the multivariate analysis for certain outcomes. Limitations included first, that specific antivirals were not named in the questionnaire. Second, that specific underlying medical conditions were not solicited nor used to define immunocompromised status. While age is the largest risk factor for severe COVID-19 outcomes, and we did have information based on self-report about people with disability and immunosuppression, we could not address knowledge, attitudes, and perceptions among people with specific medical conditions. Third, since state of disability was not asked without reference to ability to work, it is possible that any nuances among people with a disability who were able to work to some degree were not captured leading to possible under-ascertainment of disability and uncertainty in the estimates about this group’s knowledge, attitudes, and perceptions. Fourth, although we report the strongest predictors from the multivariate models, these variables accounted for less than half of the variance in each model, with some accounting for as little as 6.8% of the variance, although only predictors with at least 10% variance are highlighted. Fifth, the data were self-reported and not confirmed through medical records and vaccination records. Sixth, some responses may have been influenced by desirability bias.

## Conclusion

Among adult respondents, most respondents were knowledgeable about antivirals and agreed that they can reduce the risk for severe illness and hospitalization, and were willing to take them, particularly if recommended by a doctor. However, groups that are less likely to have been vaccinated, those living in isolation, with lower incomes, lower levels of education, and who were not sure about whether they would take an antiviral or not, may be at risk for not asking for or receiving treatment to prevent severe COVID-19 outcomes. As such they represent a potential target group who might be more open to persuasive interventions to improve their COVID-19 antiviral uptake. Understanding factors associated with current attitudes, behaviors, and willingness to use antiviral medications for COVID-19 may help identify populations and targeted policies, messages, and interventions, such as advice from a trusted provider, to bridge the gap between public knowledge and available therapies and increase COVID-19 antiviral uptake. In a pandemic such as COVID-19, synergistic functioning between public health and private and public healthcare systems may ensure that patients and health care providers are aware and knowledgeable about available treatments for COVID-19, and that patients who are eligible for these lifesaving medicines receive them. As we prepare for seasonal iterations of COVID-19 in 2024 and beyond, these results may be applicable in developing targeted messaging to improve COVID-19 antiviral uptake among those who are eligible.

## Data Availability

All data produced in the present study are available upon reasonable request to the authors

## Notes

### Competing Interest Statement

The authors have declared no competing interest.

### Funding Statement

This study was funded through a contract between the National Center for Immunizations and Respiratory Diseases Office of Communications and KRC Research

### Author Declarations

Ethics committee/IRB of the Centers for Disease Control and Prevention gave ethical approval for this work.

### Summary of Updates

Abstract results section clarified to better summarize main results; Outcomes section updated to clarify Coefficient selection cut-off and interpretation; Discussion edited to clarify how findings could be used to target messaging and interventions to potentially receptive groups and possibly improve COVID-19 antiviral use; Table 1 edited to clarify comparison groups; Tables 2a, 2b, 2c edited to clarify analytic characteristics such as model type, variables, and reference groups.

